# Evaluating the impact of non-pharmaceutical interventions for SARS-CoV-2 on a global scale

**DOI:** 10.1101/2020.07.30.20164939

**Authors:** Rachel Esra, Lise Jamieson, Matthew P. Fox, Daniel Letswalo, Nkosinathi Ngcobo, Sithabile Mngadi, Janne Estill, Gesine Meyer-Rath, Olivia Keiser

## Abstract

In the absence of a viable pharmaceutical intervention for SARS-CoV-2, governments have implemented a range of non-pharmaceutical interventions (NPIs) to curb the spread of infection of the virus and the disease caused by the virus, now known as COVID-19. Given the associated social and economic costs, it is critical to enumerate the individual impacts of NPIs to aid in decision-making moving forward. We used globally reported SARS-CoV-2 cases to fit a Bayesian model framework to estimate transmission associated with NPIs in 26 countries and 34 US states. Using a mixed effects model with country level random effects, we compared the relative impact of other NPIs to national-level household confinement measures and evaluated the impact of NPIs on the global trajectory of the COVID-19 pandemic over time. We observed heterogeneous impacts of the easing of restrictions and estimated an overall reduction in infection of 23% (95% CI: 18-27%) associated with household confinement, 10% (95% CI: 1-18%) with limits on gatherings, 12% (95% CI: 5-19%) with school closures and 17% (95% CI: 6-28%) with mask policies. We estimated a 12% (95% CI: 9-15%) reduction in transmission associated with NPIs overall. The implementation of NPIs have substantially reduced acceleration of COVID-19. At this early time point, we cannot determine the impact of the easing of restrictions and there is a need for continual assessment of context specific effectiveness of NPIs as more data become available.

## Introduction

The novel SARS-CoV-2 virus was identified in late December 2019 with first cases occurring in Wuhan City, Hubei Province, China. A rapid spread of the virus in China, followed by an exponential increase of global cases of the virus, resulted in the declaration of a global pandemic by the World Health Organization (WHO) on March 11, 2020. As of the end of July 2020, reported infections have exceeded 13 million representing a small fraction of the estimated global burden and resulting in almost than 580 000 reported deaths^1^. Transmission of SARS-CoV-2 occurs via droplets or direct contact^2^. Mortality associated with the disease cause by the virus, now known as COVID-19, is highest in those of older age and those with pre-existing vascular and respiratory conditions ^2^.

In the absence of viable pharmaceutical interventions, governments have implemented non-pharmaceutical interventions (NPIs) informed by historical outbreaks of H1N1 Influenza^3^, severe acute respiratory syndrome (SARS) and Middle East Respiratory Syndrome (MERS)^4^. Using data from control measures implemented in China in the first months of the epidemic ^5,6^, mathematical modelling studies ^7–10^, and, observational evidence ^11^, NPIs have been implemented at both national and sub-national scales, at varying times in the epidemic trajectory and at varying levels of severity. These interventions have ranged from the closure of educational institutions, social distancing measures, restrictions on public gatherings, face-mask policies and, enforcement of large-scale quarantines including stay-at-home orders, household confinements and national lockdowns. NPIs have been implemented to reduce population contact and slow down transmission rates resulting in an estimated 55% of the total global population observing some form of self-quarantine by the end of April 2020 ^12^.

Through the implementation of NPIs, a number of Asian and European countries first affected by the pandemic have curbed infection rates and have achieved epidemic control ^8^. As infection rates continue to rise in other regions, a number of European countries have begun to lift restrictions starting May 2020. A recent study utilised the incidence rate ratio, the ratio of the rate of new infections reported between two time-periods, to evaluate the impact of NPIs in 139 countries ^11^. They reported that physical distancing measures were associated with a 13% reduction of SARS-CoV-2 incidence. One of the difficulties in enumerating the quantitative impact of COVID-19 NPIs relates to the currently available measurement of transmission based on real-time reported SARS-CoV-2 incidence. Discrepancies in the coverage and strategy of testing over time, as well as delays in reporting and time to symptom onset, mean that daily-confirmed cases do not give an accurate depiction of the total infected population at a given point in time. Due to this, a number of groups have implemented dynamic mathematical modelling to estimate transmission parameters that are currently unknown ^7–10^. A recent systematic review of evidence until March 12 2020 included observational and modelling studies of SARS and MERS outbreaks and modelling studies of COVID-19 related NPIs, concluding that quarantine measures were effective in reducing transmission and COVID-19 related deaths^4^. Authors cautioned that data from SARs and MERS outbreaks may not be applicable to the current pandemic and that results from modelling studies were based on limited data at this early stage^4^.

The European Centre of Disease Prevention and Control (ECDC) developed an age-stratified dynamic transmission model to estimate the impact of NPIs in 30 European countries from the start of the pandemic to 2 May 2020^9^. In the absence of empirical evidence on the efficacy of SARS-CoV-2 related NPIs, they used expert opinion to estimate and rank the effectiveness of enforced lockdowns, voluntary home confinement, and closure of public spaces and cancellation of mass gatherings estimating an 70% median reduction in population contacts as a result of NPI implementation^9^. Using a semi-mechanistic Bayesian hierarchical model, Flaxman et al. estimated the impact of lockdowns, banning of public events, school closures, self-isolation and social distancing in eleven European countries from the start of the pandemic to May 4, 2020^8^. Due to the close and often overlapping time-periods in which interventions were implemented in the early stages of the pandemic, they were unable to ascertain the impacts of specific interventions. They did however estimate that periods of national lockdowns have led to an 81% (95% CI: 75-87%) reduction in transmission overall, as well as the prevention of 3.1 million deaths across all eleven countries ^8^. Another study [currently in preprint] utilised a Susceptible-Infected-Recovered (SIR) model with time-varying covariates for nine countries in three continents that were or currently are SARS-CoV-2 epicentres^10^. They found that travel restrictions and social distancing were not associated with reductions in transmission and instead ascertained that a combination of school closures, mask wearing and centralized quarantine may be sufficient to replace full lockdown measures^10^. Since the publication of these studies, a number of research organisations have begun to collate and validate global datasets tracking the implementation of SARS-CoV-2 NPIs by either assigning a time varying ordinal scale of the severity of interventions implemented or using a variety of intervention categorisation frameworks^13^.

To our knowledge, no-one has implemented Bayesian methodology accounting for uncertainty in SARS-CoV-2 transmission and data reporting, to quantify the impact of specific related NPIs across all global regions. Additionally, we incorporated the first global estimates of the impacts of easing of restrictions up to May 31st, 2020. Previous studies have focused on European countries first affected by the pandemic and as such the effectiveness of NPIs in other global regions where cases continue to rise is currently unknown. It is critical to enumerate the individual impacts of NPIs to aid in decision making moving forward, especially for countries in the second and third wave of affected countries. We consider the impact of the timing of the interventions in the local trajectory of the pandemic. In addition to what is currently known about the impact of NPIs in European countries in the first months of the pandemic, we include countries affected later in the global trajectory, where we expect a different impact of specific NPIs as global awareness has resulted in large-scale behavioural changes.

## Methods

### Global NPI timeline

We created a global timeline of the implementation on NPIs including quarantine and isolation policies, limits on gatherings, school closures (primary, secondary and tertiary educational institutions), mask policies, household confinements (stay-at-home-orders, shelter in place orders and lockdowns) and the easing of restrictions. We included all countries and United States (US) where COVID-19 had exceeded 5000 cases by May 31^st^, 2020. We accessed daily reported cases from the United States Centers for Disease Control (CDC) COVID Data Tracker^14^ for the US and European Centre of Disease Prevention and Control data for the rest of the world, all accessed on May 31^st^, 2020^1^. We utilised the public health and social measures global dataset curated by the WHO (WHO PHSM), a consolidated dataset of six global databases of COVID-19 related public health and social measures^13^. A full description of our validation of the dataset is detailed in the Supplementary materials. We additionally included information NPI implementation in the US from the Boston University School of Public Health US state policy database^15^. We included only compulsory NPIs implemented on a national (or state-level in US) scale and validated timelines by cross-referencing dates with official governmental and health ministry announcements. A full description of the timeline, sources and collection methodology can be found in the supplementary material. In line with methodology used in previous publications^8,9^, we assumed the effect of household confinement to supersede the effect of any other NPI implemented simultaneously and excluded any interventions implemented after household confinement and prior to easing of restrictions. We defined the start date of an intervention as the date on which the intervention was implemented and the end date as the date of any successive intervention in the timeline. Based on the assumption of a 5-day latency period for SARS-CoV-2^16^, we included all events that had been implemented for a minimum of 5 days in the absence of any other event. For interventions that are currently ongoing and where no successive interventions have been implemented, we defined the end date as the date on which the analysis was conducted (May 31st, 2020). We defined the date of easing of restrictions as the date at which any of the following was officially announced: lifting of household confinement orders, opening of schools, opening of non-essential industry activities and easing of limitations on gatherings.

### Estimation and comparison of intervention specific Rt

We utilised the EpiNow R package, developed by Abbot et al., to estimate the time varying daily reproduction numbers, Rt, from daily reported cases while accounting for uncertainty in reporting delays, incubation period and generation time^17^. A full description of the package and specifications used to propagate uncertainty are described in full elsewhere^17^. Following the methodology described, a publicly available line-list of reported cases and time to symptom onset was used to estimate uncertainty in the delay between symptom onset and case notification^18^. We estimated exponential and gamma distributions of time to symptom onsets for each confirmed case and binomial upscaling was used to account for right truncation of notification dates. This same methodology was used to estimate uncertainty in the serial interval. Assuming a normal distribution, parameters were sampled from the reporting delay distribution and serial interval distribution were generated as described elsewhere^17^. Generation time was estimated as described by Ganyani et al.^19^ using an incubation period of 5-days^16^. R0, the basic reproduction number was modelled with a gamma prior distribution using a mean estimate of 2.6 (sd: 2) based on early estimates from Wuhan^17,20^. Using the EpiEstim R package developed by Cori et al.^21^, Rt was estimated from 1000 samples at each time point using parameter specifications as described ^17^. As specified by Cori et al.^21^ the instantaneous reproduction number over time period measured by the reproduction number is distributed with a gamma distribution, i.e.

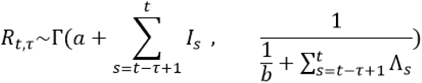

Resulting in a posterior mean equal to

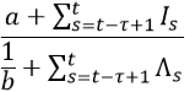

And a posterior coefficient of variation (CV, standard deviation divided by mean) equal to

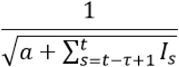

Time varying daily Rt was estimated from the day reported cases exceeded 50 to May 31^st^, 2020 (Figure 1). We calculated the average time varying daily Rt for each intervention period as well as the percentage change in daily Rt from the start to the end of each intervention period. Linear mixed effects modelling fit by restricted maximum likelihood was used to assess the relationship between the type and timing of individual NPIs and changes in Rt. We compared the relative percentage reduction in Rt associated with each NPI and the easing of restrictions by calculating the β-coefficient estimated marginal mean according to the following formula:

**Figure 1.**
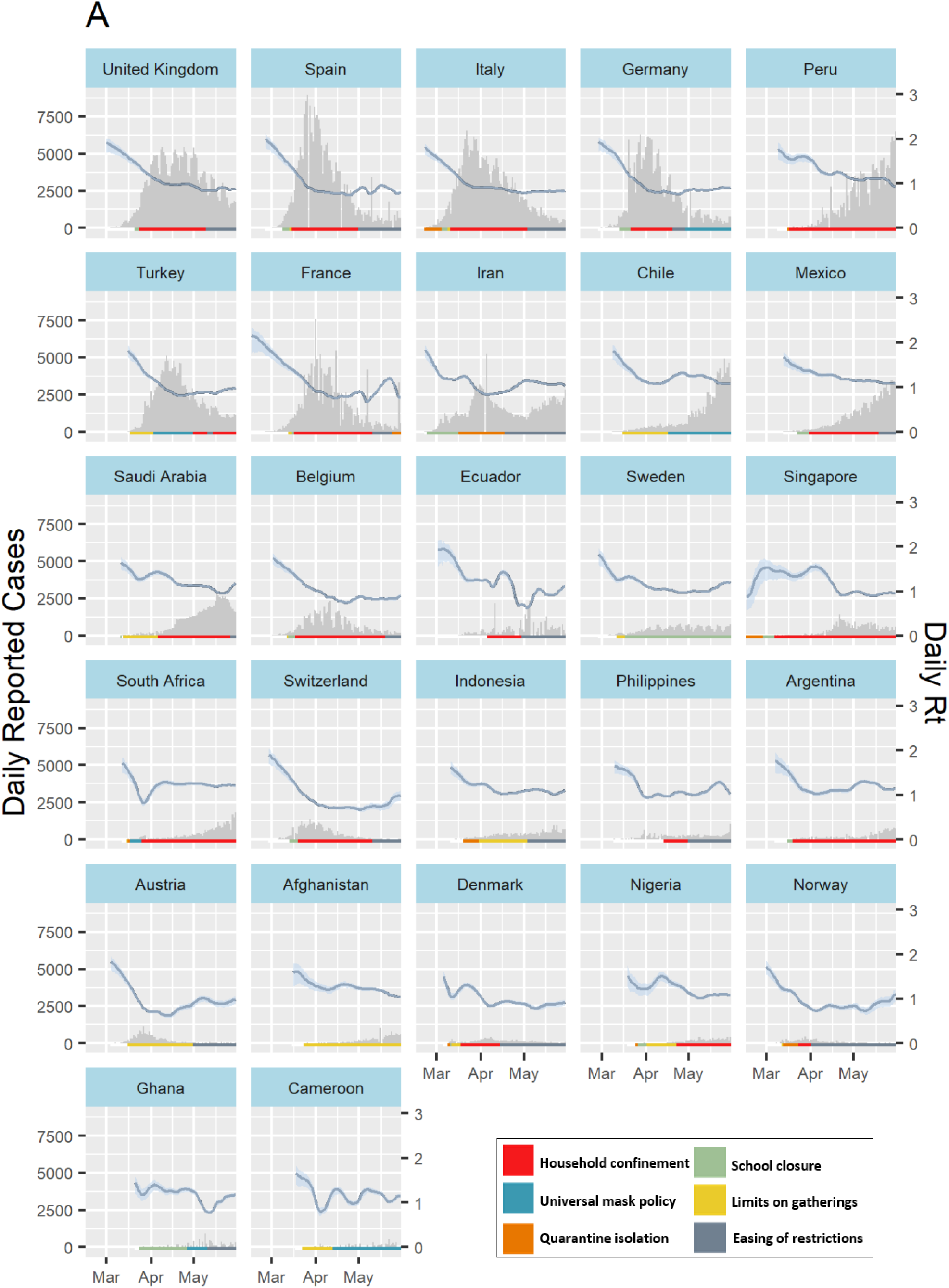
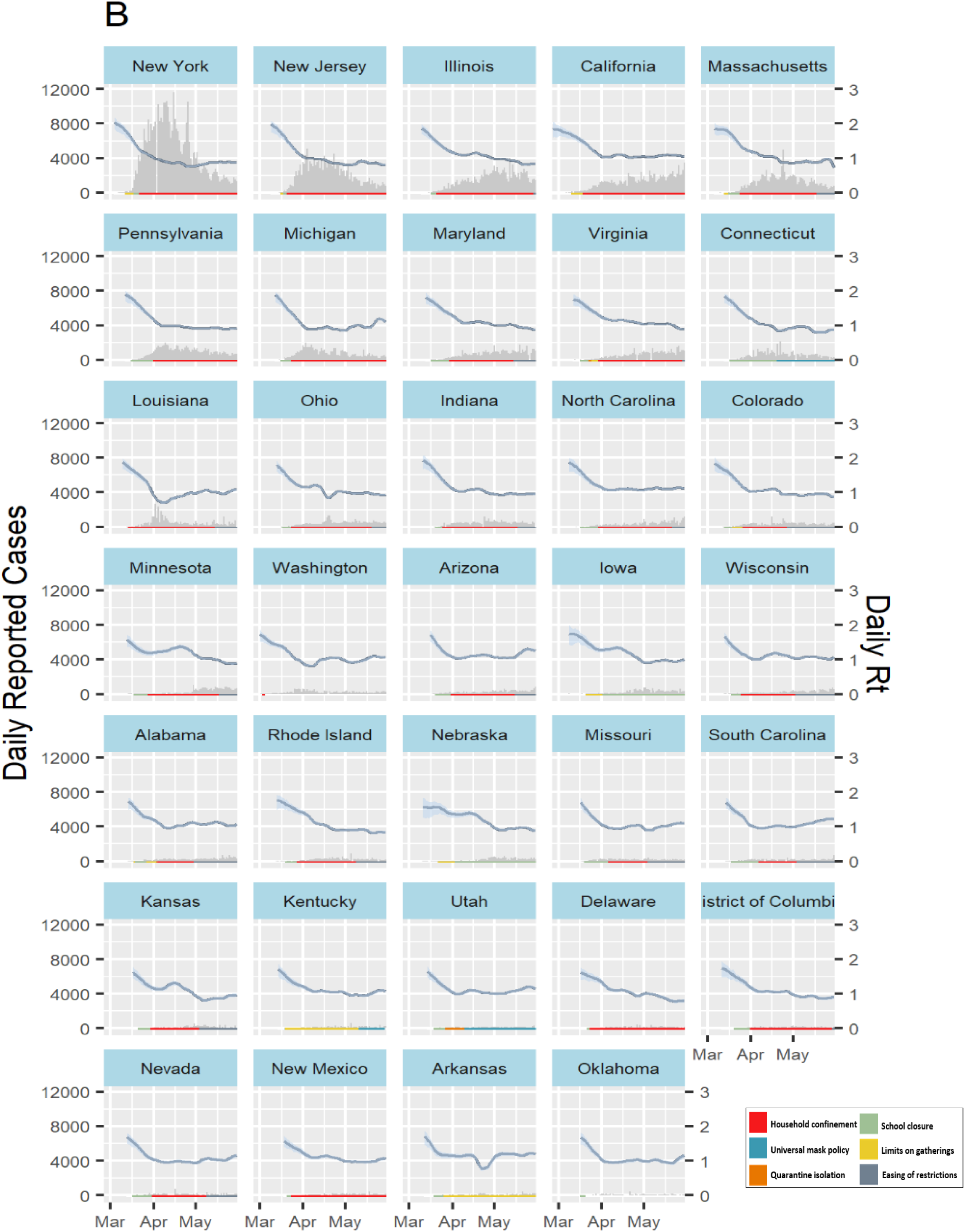
(below): Timeline of daily reported COVID-19 incidence and implementation of non-pharmaceutical interventions across different countries (A) and US states (B). Plots are arranged by order of highest cumulative caseload as of May 31st, 2020 and illustrate the timeline of the implementation of quarantine and isolation policies (orange), limits on gatherings (yellow), school closures (green) and universal facemask policies (blue). We excluded other interventions implemented during time-periods of household confinement (red) and specified the date of easing of restrictions (grey) as defined in the methods. NPI timelines are illustrated alongside daily reported cases (grey bars) and the estimated daily R_t_ (blue line and ribbon representing median R_t_ and 0.05 – 0.95 quantiles respectively).

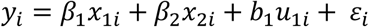

where

*y*_*i*_ is a vector of observations (% change in *R*_*t*_ over the intervention period) for country *i*

*x*_1*i*_ is the fixed-effect predictor (intervention type), with its regression coefficient *B*_1_

*x*_2*i*_ is the fixed-effect predictor (days since 50^th^ case), with its regression coefficient *B*_2_

*u*_1_ is the random-effect predictor (country), with its regression coefficient *b*_1_

*ε*_*i*_ is the error for observation in country *i*

All calculations and modelling estimations were run in R Studio using R Version 6.3.2.

## Results

### Global implementation of SARS-CoV-2 related NPIs

After the exclusion of all overlapping interventions, the final analysis consisted of 145 events from 26 countries and 34 US states (Figure 1). In the majority of scenarios, quarantine and isolation, limits on gatherings and school closures preceded the implementation of household restrictions (Figures 1 and 2). Universal facemask policies and mandatory quarantine isolation orders were often implemented in conjunction with other interventions and as a result we were only able to evaluate the impact of these interventions in 16% (N=10) and 33% (N = 21) of countries and states respectively (Table 1, Figure 1). There was large variation in the timing of the implementation of NPIs across regions (Figure 1). In Europe, where incidence has decreased in the majority of countries, 92% of countries (N = 11) have begun to ease restrictions as of May 31^st^ 2020. Comparatively, restrictions were still in place in African and South & Latin American countries where incidence is still rising. This same pattern was not observed in the United States where 71% (N = 27) of states included had begun to ease restrictions irrespective of marked decreases in incidence (Figure 1).

**Table 1:** Global coverage of the implementation and easing of Covid-19 related NPIs as of May 31^st^, 2020

|  | Africa | Middle East | Asia | Europe | South/Latin America | United States of America | Global |
| --- | --- | --- | --- | --- | --- | --- | --- |
|  | (N=3) | (N=3) | (N=3) | (N=12) | (N=5) | (N=38) | (N=64) |
| *N countries/ states implementing (% regional total)* |  |  |  |  |  |  |  |
| Quarantine isolation | 3 (100% ) | 2 (67% ) | 3 (100% ) | 6 (50% ) | 3 (60% ) | 4 (11% ) | 21 (33% ) |
| Limits on gatherings | 3 (100% ) | 2 (67% ) | 1 (33% ) | 11 (92% ) | 3 (60% ) | 24 (63% ) | 44 (69% ) |
| School closures | 3 (100% ) | 3 (100% ) | 2 (67% ) | 12 (100% ) | 5 (100% ) | 35 (92% ) | 60 (94% ) |
| Universal mask policies | 2 (67% ) | 0% ) | 0% ) | 3 (25% ) | 2 (40% ) | 3 (8% ) | 10 (16% ) |
| Household confinement | 2 (67% ) | 1 (33% ) | 2 (67% ) | 12 (100% ) | 4 (80% ) | 31 (82% ) | 52 (81% ) |
| Easing of restrictions | 1 (33% ) | 2 (67% ) | 2 (67% ) | 11 (92% ) | 2 (40% ) | 27 (71% ) | 45 (70% ) |

**Figure 2:**
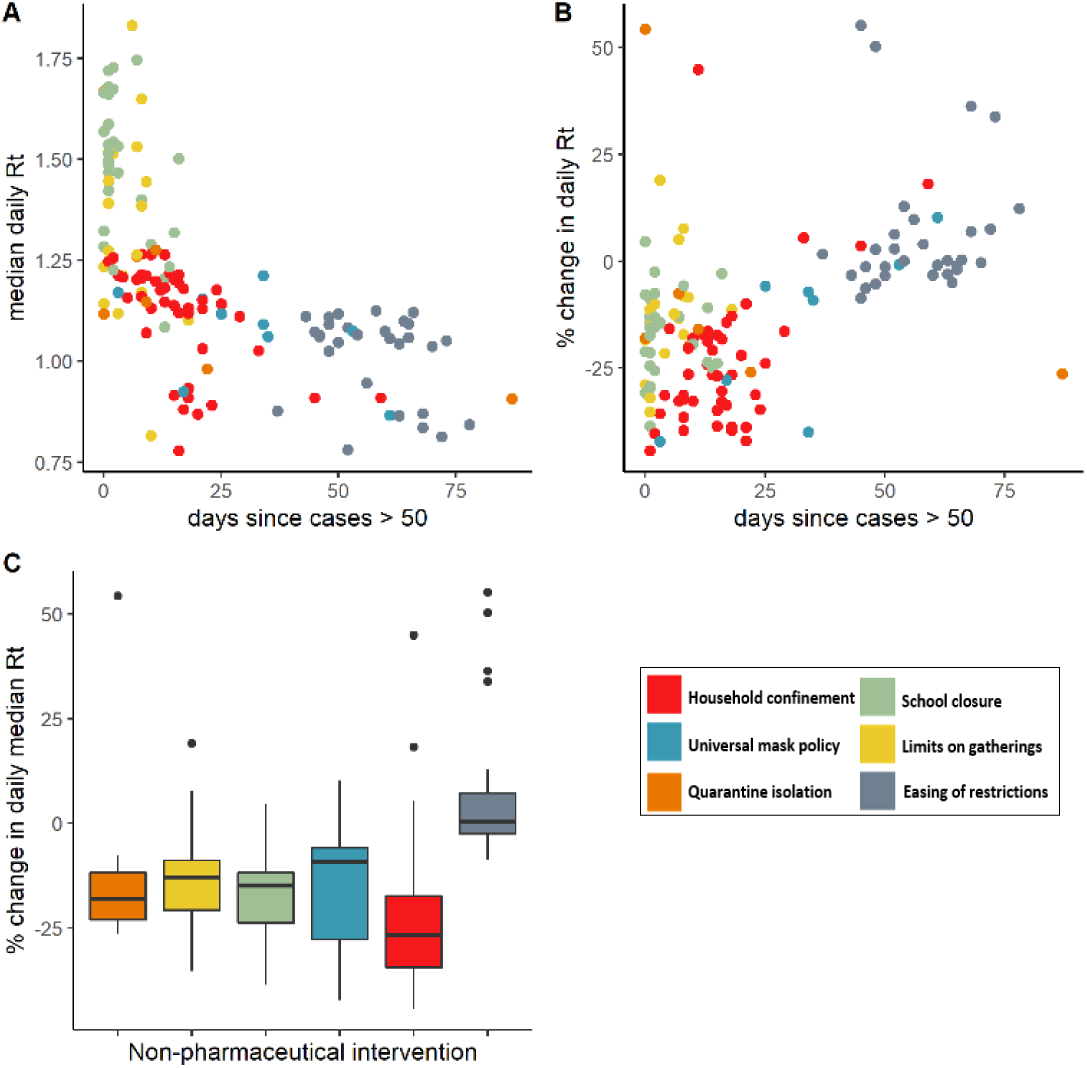
Timing and relative effect of SARS-CoV-2 related NPIs. We evaluated the median daily R_t_ **(A)** and overall change in daily R_t_ **(B)** associated with quarantine and isolation of known positives (orange), limits on gatherings (yellow), school closures (green) and universal facemask policies (blue), household confinement (red) and easing of restrictions (grey) over time. Dots represent the start date of each intervention and easing of restrictions with T=0 defined as the day since cases exceeded 50. We estimated the overall relative % in R_t_ associated with each intervention **(C)** with black lines representing the interquartile range and dots representing outlier events.

### Relative timing and effect of individual SARS-CoV-2 related NPIs

It was evident that NPIs clustered over time (Figure 2). We observed a negative correlation between mean daily Rt and relative change in Rt over time, with higher mean Rt and a larger relative reduction in Rt observed during NPIs implemented in the earlier months of the pandemic (Figure 2, A+B). We estimated overall reductions in Rt associated with household confinement (23%), limits on social gatherings (10%), school closures (12%) and mask policies (17%) (Table 2, Figure 2C). In cases where restrictions had been eased, we estimated a range of impacts on transmission with Rt both increasing and decreasing (Table 2). We additionally estimated an overall 12% (95% CI: 9-15%) reduction in daily Rt over time, irrespective of intervention implemented (Table 2). We were unable to estimate plausible estimates for country, state or regional effect sizes of the individual and overall impact NPIs.

**Table 2:** Estimated marginal means of % reduction in R_t_

| Restrictions | Reduction in $R_t$ [95% CI] |
| --- | --- |
| Household confinement | 23% [18% ; 27%] |
| Quarantine isolation | 8% [-4% ; 20%] |
| Limits on gatherings | 10% [1% ; 18%] |
| School closure | 12% [5% ; 19%] |
| Mask policy | 17% [6% ; 28%] |
| Restrictions eased | 2% [-8% ; 12%] |

In line with previous modelling studies^8,9^, we found that household confinement was associated with the largest reduction in Rt. In a time independent analysis, all other NPIs were associated with a higher Rt in comparison to household confinement as a baseline intervention (Table 3A). When adjusting for the combined impact of all NPIs (ie. the average change in Rt over time), the range of impact sizes for other NPIs widened and in all cases included zero (Table 3B), supporting the effect of time evident in the first step of our analysis (Figure 2A and B).

**Table 3:** Beta-coefficient impact estimates of % change in R_t_ associated with other NPIs, easing of restrictions and overall change in R_t_ overtime compared to household confinement

| Intervention | Beta-coefficient estimate [95% CI]* |  |
| --- | --- | --- |
|  | Time independent <sup>A</sup> | Time dependent <sup>B</sup> |
| Quarantine isolation | 0,16 [0,03 ; 0,28] | 0,15 [0,02 ; 0,27] |
| Limits on gatherings | 0,10 [0,02 ; 0,19] | 0,13 [0,04 ; 0,22] |
| School closure | 0,08 [0,00 ; 0,15] | 0,10 [0,03 ; 0,18] |
| Mask policy | 0,09 [-0,02 ; 0,21] | 0,06 [-0,06 ; 0,18] |
| Easing of restrictions | 0,30 [0,23 ; 0,38] | 0,21 [0,09 ; 0,33] |
| Change in $R_t$ over time | - | 0,002 [0,00 ; 0,05] |

\* Positive coefficients indicate an associated larger $R_t$ in comparison to household confinement
<sup>A</sup> Linear mixed effects model including only fixed effects of NPIs and easing of restrictions
<sup>B</sup> Linear mixed effects model including fixed effects of NPIs, easing of restrictions and change in $R_t$ over time

## Discussion

We evaluated the impact of specific NPIs on reported SARS-CoV-2 infection rates in a global context. We found a large variation in the timing and combination of NPIs implemented across all global regions. In 26 countries and 34 US States, we estimated a 23%, 17%, 12% and 10% reduction in the daily time varying Rt as a result of household confinement, mask policies, school closures and limits on social gatherings respectively. In agreement with current SARS-CoV-2 observational evidence^11^ and data from a recent systematic review^4^, we found that earlier implementation of NPIs had a larger impact on the reduction of Rt.. We additionally estimated an overall 12% reduction in Rt in all contexts where NPIs had been implemented. The easing of restrictions had a heterogeneous effect, owing in part to restrictions being eased in groups, making it harder to delineate the effect of lifting each NPI in turn

Globally, the majority of countries have implemented some type of NPI and we hypothesize that all have experienced large-scale behaviour changes as a result of the SARS-CoV-2 pandemic. As such we do not have a suitable counterfactual scenarios to estimate rates of infection in the total absence of NPIs.

The overall 12% reduction in Rt, over time may be representative of the overall impact of sequential interventions and general changes in behaviour ‘snowballing’ as global awareness increased, resulting in the overall reduction in Rt regardless of the type and order of interventions implemented. In line with assumptions informed by other studies^4,8^ and expert opinions^9^, we found that in the absence of any other NPI, household confinement has the largest impact on reduction in Rt. In almost all cases, additional NPIs were implemented prior to household confinements and as such, residual effects from these interventions may contribute to strong reductions observed.

We were able to estimate plausible associated reductions for the above-mentioned NPIs and found a large variation in the impact of quarantine and isolation and easing of restrictions. We reported an overall 23% (95% CI: 18-27%) reduction in Rt as a result of home confinements in comparison to the 81% (95% CI: 75-87%) estimated by Flaxman et al.^8^ The study by Flaxman et al included 11 European countries^8^, those first affected by the pandemic where the majority had reached epidemic control by the end of May 2020^7^. Our estimate of the impact was informed by the same countries as well as countries affected later on the global trajectory that have not yet reached a peak of infections and thus observed smaller reductions in R_t_ during intervention periods. Due to the large variation in impacts observed, we were unable to ascertain region and country level specific effects to further investigate this relationship.

Current observational estimates of the potential impact of NPIs on SARS-CoV-2 are based on historical interventions to control SARS and MERS outbreaks^3,4^ and the impact of NPI measured by real-time SARS-CoV-2 reports^13^. While observational evidence supports the implementation of NPIs, estimates using reported SARS-CoV-2 incidence are limited by a number of factors discussed later on. As such, dynamic mathematical models with country specific demographics and infection parameters have been used to estimate the individual impacts of NPIs on SARS-CoV-2 in China^6,22^, the UK^23^ and in Europe ^7–10^. Using a mathematical model informed by region-specific smartphone mobility data, Lai et al. estimated the role of local travel restrictions, identification and isolation of cases and social distancing measures in curbing SARS-CoV-2 infection rates in China^6^. With a detailed timeline of region specific NPI implementations and by estimating population mobility, region-specific contact patterns and resultant SARS-CoV-2 infection rates, authors concluded that lockdown alone would not have resulted in controlling infection in Wuhan if not preceded by local travel restrictions^6^. They additionally noted the importance of the timing of interventions, estimating that the implementation of NPIs a week earlier or later may have resulted in 3-fold reduction or increase in observed cases respectively^6^. Flaxman et al. evaluated the impacts of NPIs in 11 European countries, accounting for the lag in case reporting by back estimating Rt from reported death rates^8^. Due to the rapid succession of the implementation of NPIs during the beginning of the epidemic, they were unable to estimate the individual impact of NPIs with the exception of lockdown measures.

Our analysis had a number of limitations. Firstly, our metric of comparison, the time varying Rt based on reported cases, is sensitive to country level differences in testing coverage and strategies. In single country studies, it has been difficult to accurately estimate true infection rates and Rt due to the large number of asymptomatic infections that are assumed to go unreported^24^. Daily-confirmed cases do not give an accurate depiction of the total infected population at a given point in time. Country and region-specific delays in testing, in addition to differential delays between the reporting of positive and negative cases, result in a lag between daily reported cases and the actual burden of infection. A combination of rapidly evolving testing recommendations^25,26^, variations in laboratory processing capacities and, reagent shortages have resulted in the implementation of a number of different testing strategies across countries and over time. It is uncertain whether mass testing, shown to successful in the case of South Korea^27^, is feasible in other contexts, and modelling studies indicate that in the absence of contact tracing, widespread testing will not be sufficient in containing infection rates^27,28^.

Secondly, this study was limited by the epidemiological complexity of ascertaining the impacts of individual COVID-19 NPIs implemented thus far. NPIs have been implemented with overlapping timeframes, at varying levels of severity and have been adhered to with differing levels of compliance, even within the same country. In order to account for this, we evaluated only interventions that had been implemented in absence of any other intervention for a minimum period of five days. This criterion was based on the conservative assumption of a 5-day latency period for SARS-CoV-2, although reports estimate that an individual may be infectious for anywhere from 3-14 days^16^. Based on this, it may take up to two weeks to observe the impact of any given intervention (or even longer taking reporting delays into account), and therefore it is difficult to disentangle the immediate effect of an intervention and the residual impact of an intervention implemented previously. In order to account for this and the above-mentioned limitations of reported case data, we used a Bayesian methodology to model Rt with uncertainty in time to symptoms onset, reporting delays and a range of estimates for the serial interval and generation time based on currently available reports. These relationships are further confounded by static population-specific factors such as age distributions, population density and distribution of underlying comorbidities that would contribute to a differential rate of infections between countries. In addition to time-varying factors such as changes in population mobility and case-importation rates that are not well documented, interventions themselves were implemented at different time points in the global trajectory of the pandemic.

In agreement with what has been previously published^4,7–11^, we find that the implementation of non-pharmaceutical interventions (NPIs) has been effective in reducing the acceleration of the COVID-19 pandemic on a global level and that earlier implementation of NPIs results in stronger reduction in transmission. In comparison to studies based in European countries, we estimate a more conservative overall impact of lockdown measures, likely driven by the inclusion of additional countries affected later in the global trajectory of the epidemic. We find that this effect remains after controlling for time since first 100 infections. At this early stage we have been unable to quantify the impact of easing of SARS-CoV-2 restrictions or determine whether NPI implementation will result in epidemic control in countries where cases are still rising. There is a need for continual assessment of the efficacy and optimal combination of individual NPIs as more data become available.

## Data Availability

Source code is available at https://github.com/epiforecasts/covid, unless otherwise noted.

https://gitlab.com/igh-idmm-public/covid-19/covid19_npi_globalrt_estimations

## Author contributions

R.E., L.J. and G.M. conceptualised this research, R.E. conducted data analysis and visualization, R.E., D.L, N.N. and S.M compiled the intervention timeline, R.E. wrote the original draft, L.J., G.M., M. F., J.E. and O.K. reviewed and edited the final draft.

## Competing interest

The author(s) declare no competing interests.

## Data availability

Code and data used for this analysis are open source and can be found here: https://gitlab.com/igh-idmm-public/covid-19/covid19_npi_globalrt_estimations

## Role of the funding source

Rachel Esra is supported in part by the European Union’s Horizon 2020 research and innovation programme and the Swiss Excellence Foreign Scholar Grant. Olivia Keiser is supported by the Swiss National Science Foundation Grants 163878 and 196270.This study has additionally been made possible by the generous support of the American People and the President’s Emergency Plan for AIDS Relief (PEPFAR) through USAID under the terms of Cooperative Agreement 72067419CA00004 to HE2RO. The contents are the responsibility of the authors and do not necessarily reflect the views of PEPFAR, USAID or the United States Government.

## Notes

### Competing Interest Statement

The authors have declared no competing interest.

### Author Declarations

This research did not involve human or animal subjects, and ethical approval was therefore not required

